# Verbal Communication using Intracortical Signals in a Completely Locked In-Patient

**DOI:** 10.1101/2020.06.10.20122408

**Authors:** Ujwal Chaudhary, Ioannis Vlachos, Jonas B. Zimmermann, Arnau Espinosa, Alessandro Tonin, Andres Jaramillo-Gonzalez, Majid Khalili-Ardali, Helge Topka, Jens Lehmberg, Gerhard M. Friehs, Alain Woodtli, John P. Donoghue, Niels Birbaumer

## Abstract

Patients with amyotrophic lateral sclerosis (ALS) can lose all muscle-based routes of communication as motor neuron degeneration progresses, and ultimately, they may be left without any means of communication. While others have evaluated communication in people with remaining muscle control, to the best of our knowledge, it is not known whether neural-based communication remains possible in a completely locked-in state (CLIS). With consent and approval of the participant, family, and legal authority for this study, two 64 microelectrode arrays were implanted in the supplementary and primary motor cortex of a CLIS patient with ALS. The patient modulated neural firing rates based on auditory feedback, and he used this strategy to select letters one at a time to form words and phrases to communicate his needs and experiences. This case study provides novel evidence that brain-based volitional communication is possible even in a completely locked-in state.

## Introduction

Amyotrophic lateral sclerosis (ALS) is a devastating neurodegenerative disorder that leads to the progressive loss of voluntary muscular function of the body^1^. As the disorder typically progresses, the affected individual loses the ability to breathe due to diaphragm paralysis. Upon accepting artificial ventilation and with oro-facial muscle paralysis, the individual in most cases can no longer speak and becomes dependent on assistive and augmentative communication (AAC) devices^2,3^, and may progress into the locked-in state (LIS) with intact eye-movement or gaze control^4,5^. Several invasive^6-10^ and non-invasive^11-16^ brain-computer interfaces (BCIs) have provided communication to individuals in LIS^17-20^ using control of remaining eye-movement or (facial) muscles or neural signals. Once the affected individual loses this control to communicate reliably, no existing assistive technology has provided voluntary communication in this completely locked-in state (CLIS)^17-20^. Non-invasive^11-16^ and invasive^6-10^ BCIs developed for communication have demonstrated successful cursor control and sentence formation by individuals up to the stage of LIS. However, none of these studies has demonstrated communication at the level of voluntary sentence formation in CLIS individuals without stable and reliable eye-movement/muscle control, leaving the possibility open that once all movement - and hence all possibility for communication - is lost, neural mechanisms to produce communication will concurrently fail.

Here, we established that an individual was in the CLIS state and demonstrated that sentence-level communication is possible without relying upon vision. This individual lacked reliable voluntary eye-movement and, consequently, was unable to use an eye-tracker for communication. He also showed increasingly unstable and ultimately failing performance with a non-invasive eye-movement-based communication system. To restore communication in CLIS, this participant was implanted with intracortical microelectrode arrays in two motor cortex areas. The patient then employed an auditory-guided neurofeedback-based strategy to modulate neural firing rates to select letters and to form words and sentences using custom software. This individual, who before implantation was unable to express his needs and wishes through non-invasive methods, including eye-tracking, visible categorization of eye-movements, or an eye movement-based BCI-system, started using the intracortical BCI system for voluntary verbal communication three months after implantation.

## Materials and Methods

The medical procedure was approved by the Bundesinstitut für Arzneimittel und Medizinprodukte (“BfArM”, The German Federal Institute for Drugs and Medical Devices). The study was declared as a Single Case Study and has received a special authorization (“Sonderzulassung”) by BfArM, according to §11 of the German Medical Device Law (“Medizin-Produkte-Gesetz”) on December 20, 2018, with Case Nr. 5640-S-036/18. The Ethical Committee of the Medical Faculty of the Technische Universität München Rechts der Isar provided support to the study on 19 Jan 2019, along with the explicit permission to publish on 17 February 2020. Before the patient transitioned into CLIS, he gave informed consent to the surgical procedure using his eye movements for confirmation. The patient was visited at home by authors HT and JL, and thorough discussions were held with the legally responsible family members in order to establish convincing evidence of the patient’s informed consent and firm wish to undergo the procedure. The legally responsible family members then provided written permission to the implantation and the use of photographs, videos, and portions of his protected health information to be published for scientific and educational purposes. In addition, a family judge at patient’s home county court gave the permission to proceed with the implantation after reviewing the documented consent and a visit to the patient.

### Patient

The patient in mid 30’s was diagnosed with progressive muscle atrophy, a clinical variant of non-bulbar ALS, selectively affecting spinal motor neurons approximately 3.5 years before the implantation. He lost verbal communication and the capability to walk 3 years before the implantattion. He has been fed through a percutaneous endoscopic gastrostomy tube and artificially ventilated 2.5 years before the implantation and is in home care. He started using the MyTobii eye-tracking-based assistive and augmentative communication (AAC) device 2.5 years before the implantation. Aprroximately 2 years before the implantation, he could not use the eye-tracker for communication because of his inability to fixate his gaze. Subsequently, the family developed their own paper-based spelling system to communicate with the patient by observing the individual’s eye movements. According to their scheme, any visible eye movement was identified as a “yes” response, lack of eye movement as “no”. The patient anticipated complete loss of eye control and asked for a new communication system, which motivated the family to contact authors NB and UC for alternative approaches. Initial assessment sessions were performed one year before the implantationin. During this interval, the detection of eye movements by relatives became increasingly difficult, and errors made communication attempts impossible up to the point when communication attempts were abandoned. The patient and family were informed that a BCI-system based on electrooculogram (EOG) and/or electroencephalogram (EEG) might allow “yes” - “no” communication for a limited period.

The patient began to use the non-invasive eye movement-based BCI-system described in Tonin et al.^21^. The Patient was instructed to move the eyes (“eye-movement”) to say “yes” and not to move the eyes (“no eye-movement”) to say “no”. Features of the EOG signal corresponding to “eye-movement” and “no eye-movement” or “yes” and “no” were extracted to train a binary support vector machine (SVM) to identify “yes” and “no” response. This “yes” and “no” response was then used by the patient to auditorily select letters to form words and hence sentences. The patient and family were also informed that non-invasive BCI-systems might stop functioning satisfactorily, and in particular, selection of letters might not be possible if he became completely locked-in (where no eye movements could be recorded reliably). In that case, implantation of an intracortical BCI-system using neural spike-based recordings might allow for voluntary communication. As the patient’s ability to communicate via non-invasive BCI systems deteriorated, in mid of 2018, preparations for the implantation of an intracortical BCI system were initiated. To this end, HT and JL and GF were approached in order to prepare the surgical procedure and ensure clinical care in a hospital close to the patient’s home. The patient was able to use the non-invasive BCI system employing eye-movement to select letters, words, and sentences until beginning of 2019, as described in Tonin et al. ^21^. By the time of implantation, the EOG/EEG based BCI system failed, as signals could not be used reliably for any form of communication in this investigational setting. The EOG/EEG recordings and their analysis are described in Supplementary text 1 and Supplementary Figure S2. Additionally, the patient reported low visual acuity caused by the drying of the cornea.

#### Surgical Procedure

A head MRI scan was performed to aid surgical planning for electrode array placement. The MRI scan did not reveal any significant structural abnormalities, in particular no brain atrophy or signs of neural degeneration. A neuronavigation system (Brainlab, Munich, Germany) was used to plan and perform the surgery. In the first quarter of 2019, two microelectrode arrays (8×8 electrodes each, 1.5mm length, 0.4mm electrode pitch; Blackrock Microsystems LLC) were implanted in the dominant left motor cortex under general anesthesia. After a left central and precentral trephination, the implantation sites were identified by neuronavigation and anatomical landmarks of the brain surface. A pneumatic inserter was used (Hochberg et al. 2006) ^22^ to insert the electrode arrays through the arachnoid mater, where there were no major blood vessels. The pedestal connected to the microelectrode arrays connected via a bundle of fine wires (Blackrock Microsystems LLC), was attached to the calvaria using bone screws and was exited through the skin. The first array was inserted into the hand area region of the primary motor cortex^23^, and the second array was placed 2cm anteromedially from the first array into the region of the supplementary motor area (SMA) as anatomically identified. No implant-related medical adverse events were observed. After three days of post-operative recovery, the patient was discharged to his home.

##### Neural Signal Processing

A digitizing headstage and a Neural Signal Processor (CerePlex E and NSP, Blackrock Microsystems LLC) were used to record and process neural signals. Raw signals sampled at 30kS/s per channel were bandpass filtered with a window of 250-7500 Hz. Single and multi-unit action potentials were extracted from each channel by identifying threshold crossings (4.5 times root-mean-square of each channel’s values). Depending on the activity and noise level, thresholds were manually adjusted for those channels used in the BCI sessions after visual inspection of the data to exclude noise but capture all of the visible spikes above the threshold. Neural data were further processed on a separate computer using a modified version of the CereLink library (provided by Blackrock Microsystems LLC) and additional custom software. For communication, we used spike rates from one or more channels. A spike rate metric (SRM) was calculated for each channel by counting threshold crossings in 50 ms bins. The SRM was calculated as the mean of these bins over the past one second.

###### Neurofeedback communication

The patient was provided auditory feedback of neural activity levels by mapping the SRM for one or more channels to the frequency of an auditory feedback tone, as shown in Figure 1. Single channel spike rates were normalized according to the spike rate distribution of each channel. Selected channels’ normalized SRMs were then summed and linearly mapped to the range of 120-480 Hz, determining the frequency of the feedback tone produced by an audio speaker. Feedback tones were updated every 250ms. The firing rate *r*_*i*_ of each selected channel was constrained to the range [*a*_*i*_, *b*_*i*_], normalized to the interval [0,1], and optionally inverted, and the resulting rates were averaged:

**Figure 1:**
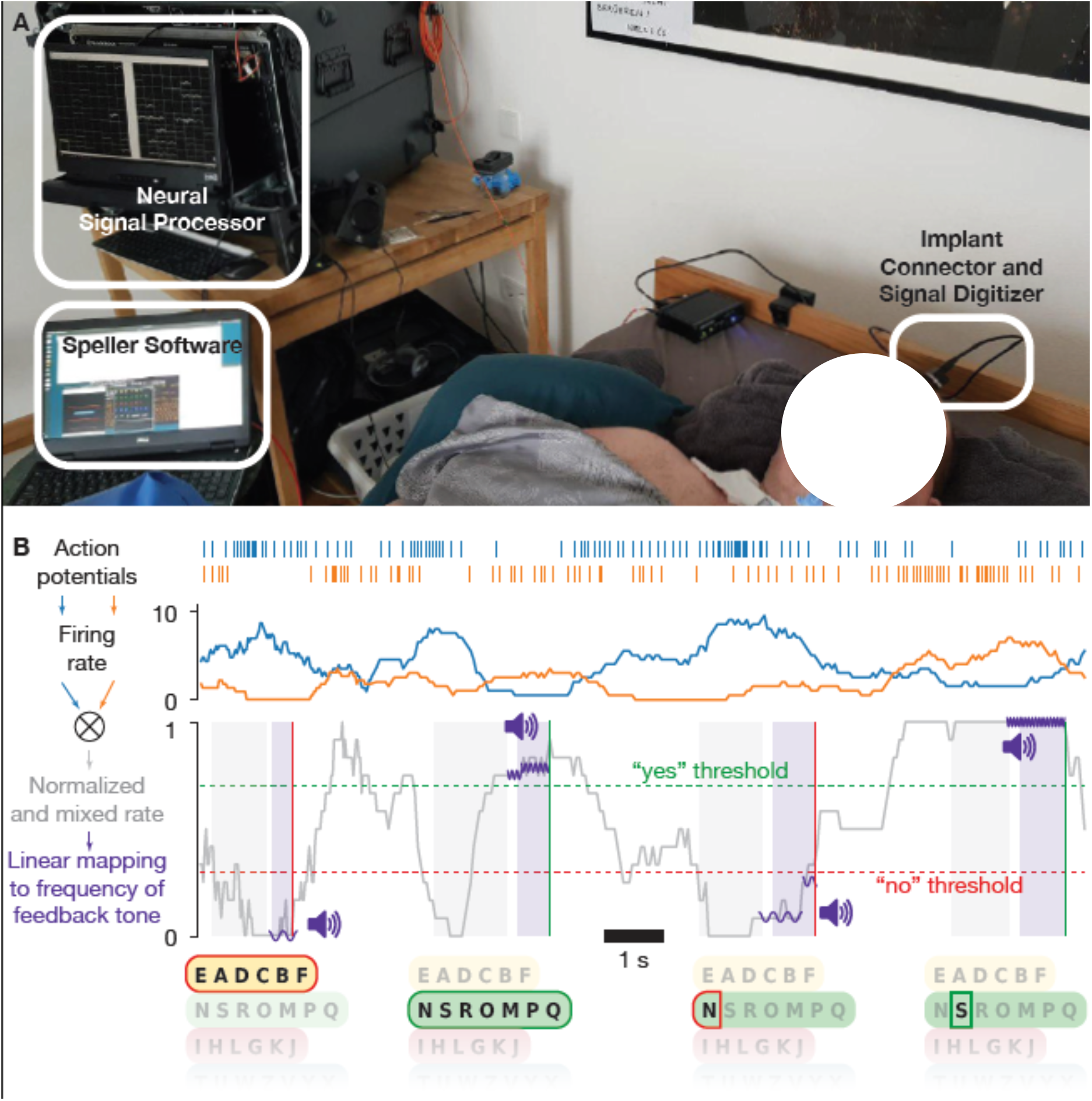
Intracortical auditory speller-based communication. A) Setup at the patient’s home. Signals were recorded from microelectrode arrays implanted in the motor cortex and processed with custom BCI software. B) Schematic representation of auditory neurofeedback and speller. Action potentials were detected and used to estimate neural firing rates. One or several channels were selected, their firing rates normalized and mixed (see Online Methods). Options such as letter groups and letters were presented by a synthesized voice, followed by a response period during which the patient was asked to modulate the normalized and mixed firing rate up for a positive response and down for a negative response. The normalized rate was linearly mapped to the frequency of short tones that were played during the response period, to give feedback to the patient. The patient had to hold the firing rate above (below) a certain threshold for typically 500ms to evoke a “Yes” (“No”) response. Control over the neural firing rates was trained in neurofeedback blocks, in which the patient was instructed to match the frequency of target tones.

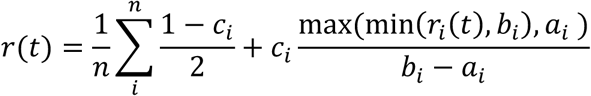

where *r*(*t*) is the overall normalized firing rate, and the *c*_*i*_ are 1 or −1. The normalized rate was then linearly mapped to a frequency between 120 and 480 Hz for auditory feedback. Feedback tones were pure sine waves lasting 250 ms each. Initially, channels were selected randomly for feedback. Then the parameters *a*_*i*_, *b*_*i*_, *c*_*i*_ as well as the channels used for control were chosen and iteratively optimized each day in the neurofeedback training paradigms.

The first paradigm (“feedback without reward”) provided successive target tones at 120 or 480 Hz, and the patient was asked to match the frequency of the feedback to the target (typically 20 pseudorandom trials per block). In the “feedback with reward” paradigm, was essentially the same, however, upon reaching and holding (during a configurable number of interactions, each interaction lasting 50ms) the feedback tone within a predefined range around the target frequency, an additional reward sound was delivered for 250ms indicating successful performance to the patient. Holding the feedback tone at the high (low) end of the range for 250ms was then interpreted by the patient upon instruction as a Yes (No) response. The “feedback with reward” paradigm served to train and validate the responses.

We also validated the Yes/No responses in a question paradigm, in which the answers were assumed to be known to the patient. Furthermore, we used an ‘exploration’ paradigm to test if the patient’s attempted or imagined movements could lead to modulation of firing rates.

Finally, in an auditory speller paradigm, the patient could select letters and words using the previously trained Yes/No approach. The auditory speller paradigm is depicted in Figure 1C. The speller system described here avoids long adaptation and learning phases because it is identical to the one used previously when he was still in control of eye movements.

The speller’s output was rated for intelligibility by three of the authors (UC, IV, and JZ). Three categories were used: unintelligible, ambiguous, and intelligible. Ambiguous speller output includes grammatically correct words that could not be interpreted in the context as well as strings of letters that could give rise to uncertain interpretations. Intelligible phrases may contain words with spelling mistakes or incomplete words, but the family or experimenter identified and agreed upon their meaning.

## Results

One day after the implantation, attempts were initiated to establish communication. The patient was asked to use his previous communication strategy employing eye movements and later to attempt hand, tongue, or foot movements to elicit neural responses. No qualitative differential neural responses could be measured. Subsequently, the communication strategy was changed on day 86, and neurofeedback-based paradigms (described in the Online Methods section) were employed, as shown in Figure 1. The patient was provided auditory feedback of neural activity by mapping a spike rate metric (SRM) for one or more channels to the frequency of an auditory feedback tone, as displayed in Figure 1 (described in the “Neurofeedback communication” section of Online Methods). The patient was able to modulate the sound tone immediately on day 86 and subsequently was able to successfully modulate the neural firing rate and match the frequency of the feedback to the target for the first time 98 days after implantation. Employing the neurofeedback strategy, the patient was able to modulate the neural firing rate and was able to use this method to select letters and to free spell from day 106 onwards. Data reported here include data from days 106–247 after implantation. During this period, the patient was visited on 47 days. Each day, we performed several neurofeedback blocks, each typically consisting of 10 high-frequency target tones and 10 low-frequency target tones presented in pseudo-random order to tune and validate the classifier. If the patient was able to match the frequency of the feedback to the target in 80% of the trials, we proceeded with the speller. Figure 2 shows the amount of time the patient was exposed to different paradigms.

**Figure 2:**
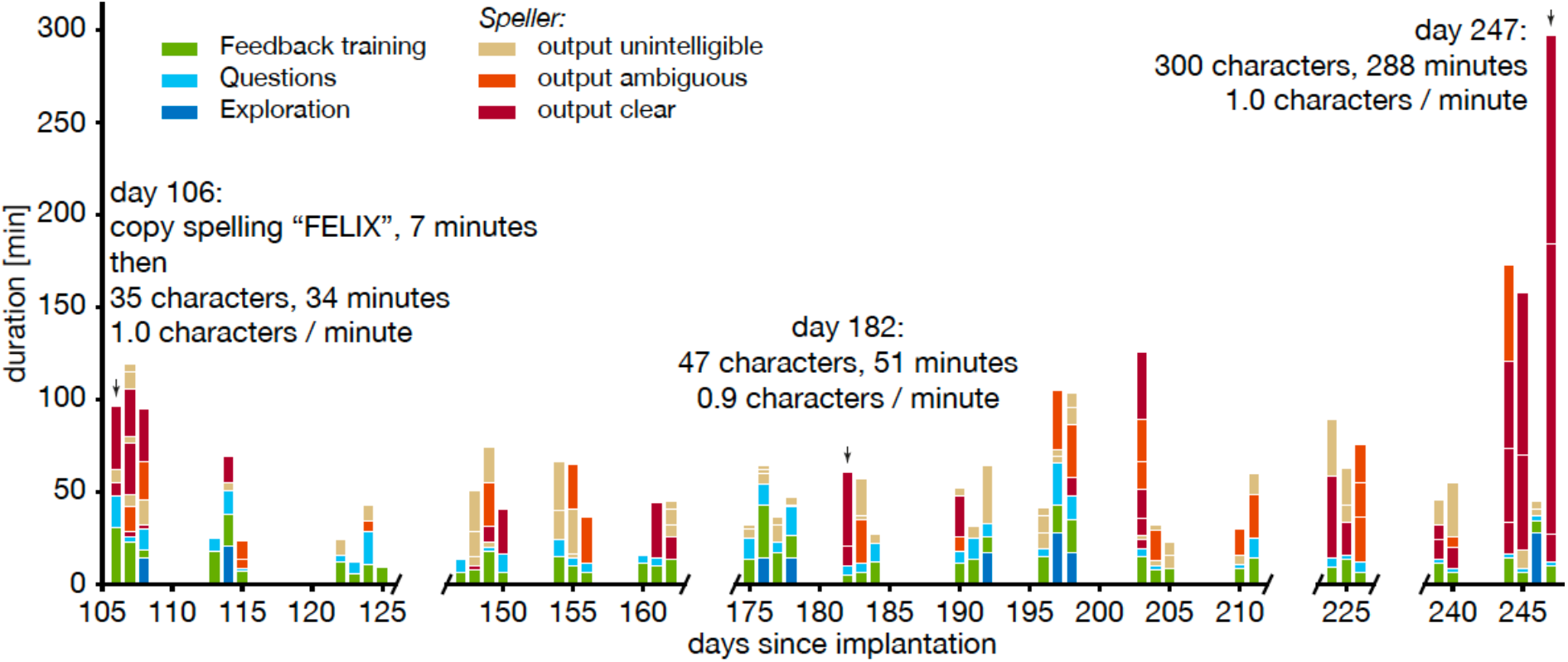
Overview of BCI use. Time the patient spent in BCI paradigms. Individual speller blocks are shown in their respective order, times for other kinds of blocks are summed per day. The x-axis represents the number of days after implantation and the y-axis the duration of the respective paradigm in minutes. Different colors represent different paradigms, as described in the legend on the top left corner of the figure. The patient was visited 3 to 4 days per week, as shown by 3 to 4 consecutive bars. The patient was hospitalised due to unrelated adverse events between days 120 and 145, 163 and 172, and 212 and 223. Question and exploration tasks are not further discussed in this paper.

Figure 3A shows individual neurofeedback trials, including an error trial, of one representative block. In the 85 neurofeedback blocks preceding the speller blocks, 1507 of 1700 trials (89%) were correct (Figure 3B), i.e., for target tone up (higher frequency) the decision was up (a “yes” answer), and for target down (low frequency) the decision was down (a “no” answer). The difference in error rates between ‘up’ and ‘down’ trials, i.e., the fraction of trials in which the modulated tone did not match the target tone, (13% and 8%, respectively), was significant (Pearson’s χ^2^ test: p<0.01). The patient maintained a high level of accuracy in the neurofeedback condition throughout the reported period: on 74% of the days, the accuracy was at least 90% during at least one of the feedback trial blocks, i.e., the patient was able to match the frequency of the feedback to the target 18 out of 20 times. We observed considerable within-day variability of neural firing rates and hence performance of the neurofeedback classifier, necessitating manual recalibration throughout the day (see Supplementary Figure S1).

**Figure 3:**
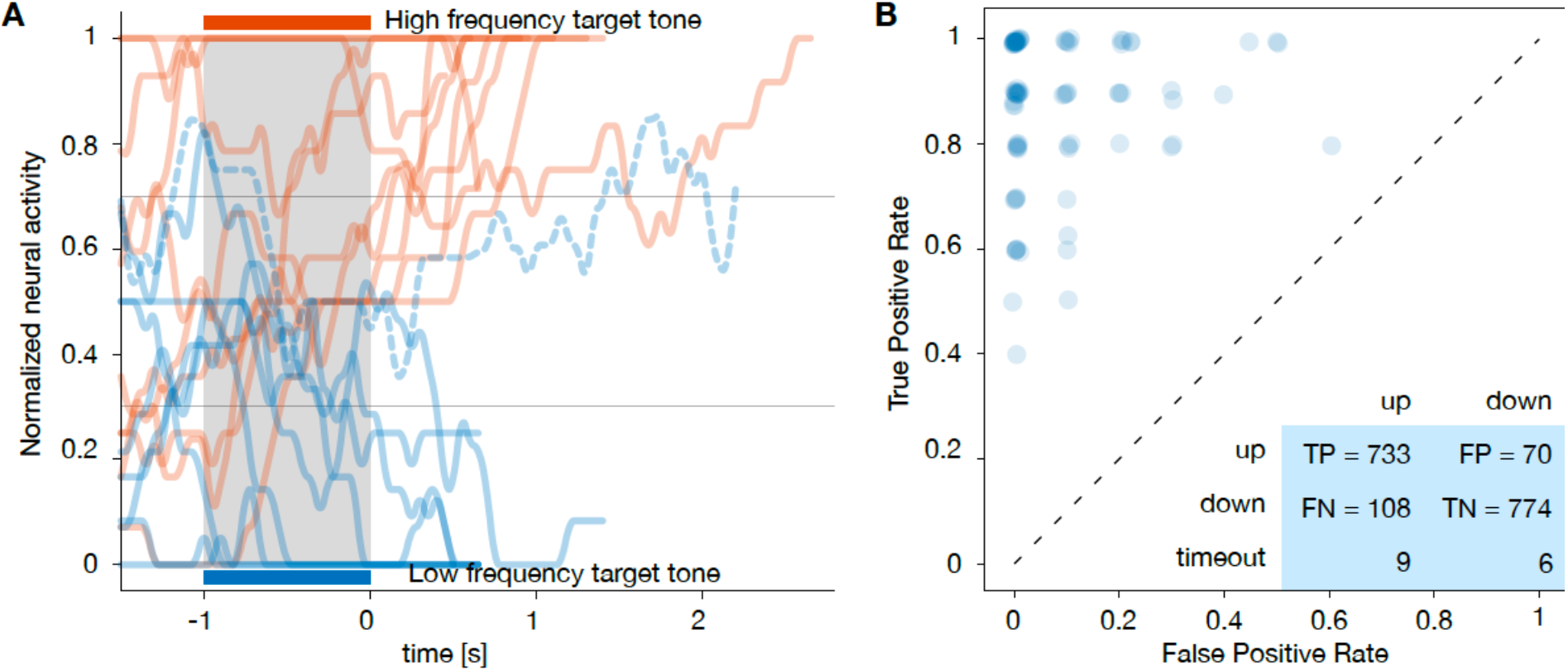
Neurofeedback task and classification. A) Representative example for normalized and mixed firing rate during ten high (red) and ten low (blue) frequency trials. The patient was asked to match the target tone by modulating the normalized and mixed firing rate, and he succeeded in all, but one trials. Trials were completed as soon as the firing rate was held above (below) the upper (lower) threshold. These blocks were used for parameter tuning, training, and validation, and performed during every day of recordings. B) True positive rate vs. false positive rate of the trials in auditory neurofeedback blocks directly preceding speller blocks on days 123–247. Each circle represents one neurofeedback block; circles are jittered for better visibility. The insert shows a contingency table of all trials in these blocks. Blocks on days 106– 122 were excluded from this analysis because time-out and error trials were treated differently in these blocks.

We continued with the speller paradigm when the patient’s performance in a neurofeedback block exceeded an acceptance threshold (usually 80%). To verify that good performance in the neurofeedback task translated to volitional speller control (based on correct word spelling), we asked the patient to copy words before allowing free spelling. On the first three days of speller use, the patient correctly spelled the specified words. After an unrelated stay at the hospital, we again attempted the speller using the same strategy on day 148.

Afterward, we relied on a good performance in the neurofeedback task, i.e., the patient’s ability to match the frequency of the feedback to the target in 80% of trials, to advance to free spelling. The selection of two letters from a speller block on day 108 is shown in Figure 4.

**Figure 4:**
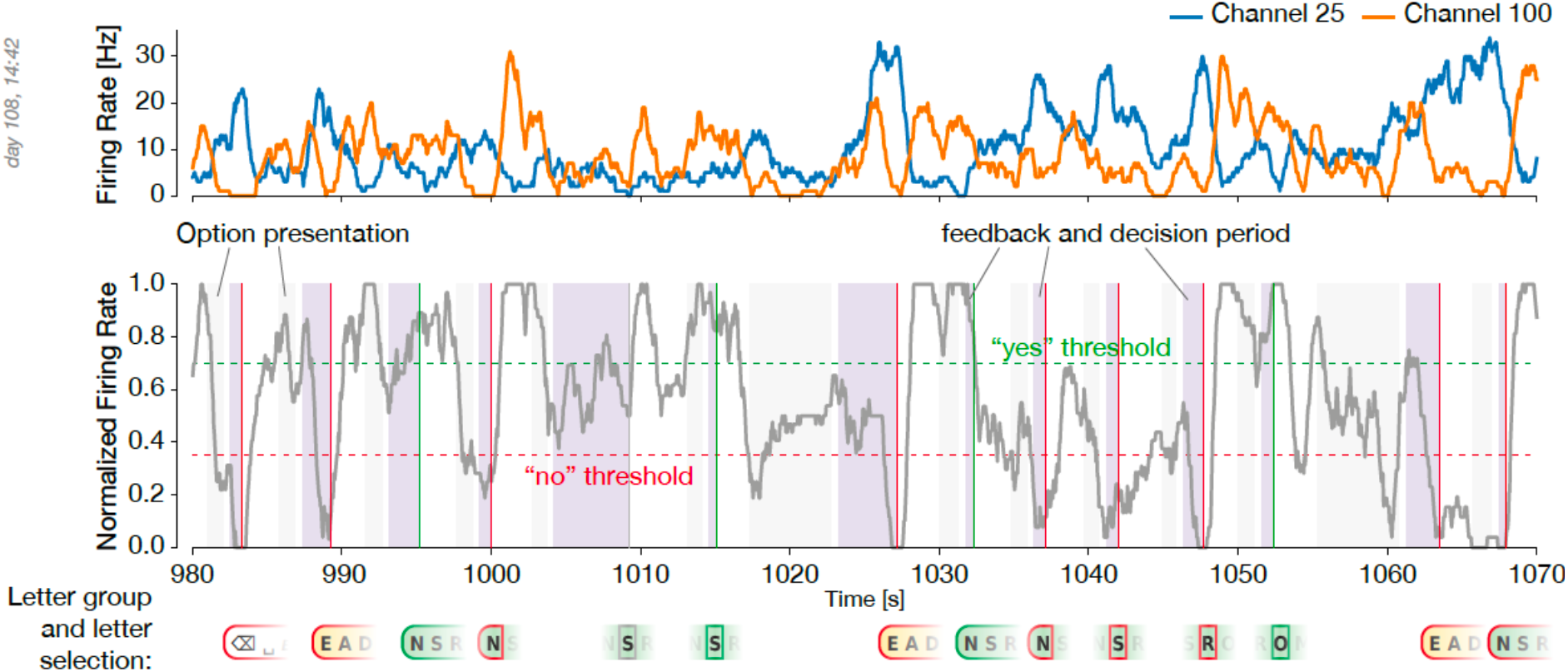
Example of letter selection during a free spelling block. A) Firing rate of the channels used for “yes”/”no” classification and B) normalized firing rate during the same 90 second period of a speller block. “Yes” / “no” / timeout decisions are marked by vertical lines. Below these lines, the corresponding selection of letter groups/letters is shown. This example is part of the phrase “DEKUBITUS PO ER **SO**LL ARME MAXIMAL”, referring to bedsore and instructing the aide to change arm position.

The patient produced comprehensible output, as rated independently by three observers, on 20 of 42 days when the speller was used (Figure 2). The median length of these communications was 26.5 characters per day, and the patient’s intelligible messages comprised 919 characters produced over 994 minutes, corresponding to an average rate of 0.92 characters per minute. This rate varied across blocks (min/median/max: 0.4/1.1/4.3 characters per minute). Over the reported period, there was no apparent trend in spelling speed.

Many of the patient’s communications concerned his care (e.g. ‘KOP?F IMMERLQZ GERAD’ – ‘head always straight’, day 161; ‘KEIN SHIRT ABER SOCKEN’ – ‘no shirts but socks [for the night]’, day 244). The patient also participated in social interactions and asked for entertainment (‘COME TONIGHT [to continue with the speller]’, day 203, ‘WILI CH TOOL BALBUM MAL LAUT HOERENZN’ – ‘I would like to listen to the album by Tool [a band] loud’, day 245, ‘UND JETWZT EIN BIER’ – ‘and now a beer’, day 247). He even gave suggestions to improve his speller performance by spelling ‘TURN ON WORD RECOGNITION’ on day 183 in English as the experimenter UC mostly spoke in English with the patient. While the speller software offered the option to delete misspelled characters, the patient often chose to continue spelling when the intended meaning was apparent, possibly because error correction is slow and time-consuming.

## Discussion and Conclusion

We demonstrate that a paralyzed patient, according to the presently available physiological and clinical criteria in the completely locked-in state (CLIS), could volitionally select letters to form words and phrases to express his desires and experiences using a neurally-based neurofeedback system. The patient used this intracortical BCI based on voluntarily modulated neural spiking from the motor cortex to spell semantically correct and personally useful phrases. In all blocks, measurable spike rate differentiation between “yes” and “no” during the neurofeedback trials and “select” and “no selection” during speller blocks appeared in few (one to five) channels in SMA (supplementary motor area) out of all active channels, which varied between 40 and 60 channels in total over the sessions. Consequently, typically only one or two channels were used for control. Spelling speed was highly variable, ranging from a few minutes to an hour to spell a short phrase. Free voluntary spelling mainly concerned requests related to body position, health status, pain, and social activities.

It is worth mentioning that no universally accepted clinical definition exists to distinguish LIS from CLIS; the current standard criteria to differentiate LIS from CLIS is the presence or absence of means of communication. During the transition from LIS to CLIS, patients are first left with limited, and finally, no means of communication. The time course of this transition process is patient and disease-specific. In theory, other voluntary muscles than eye-movements could have been used for Electromyography (EMG)-based communication attempts. Particularly face muscles outside the extraocular muscles may remain under voluntary control in some cases even after the loss of eye-muscle control. In the case of this patient, extensive EOG recordings were performed to demonstrate that no other neuromuscular output existed. The patient employed an EOG-based BCI for communication successfully for the last time in February 2019.

Nevertheless, extensive post-hoc EOG analysis showed a significant difference in the maximum, mean, and variance feature of the eye movement corresponding to “yes” and “no” even after the patient’s inability to employ the EOG-based system for communication. This failure to communicate despite the presence of a significant difference in some of the features may be due to the limitations of the EOG-based BCI system. However, such limitations cannot be answered with certainty with the post-hoc analysis used here. In this study, caretakers and family members denied and lamented any reliable communication possibility from February 2019 onwards. Thus, we conclude based on our reported measurements that the patient described was in a CLIS a few weeks before and as well as after implantation. This statement does not exclude the possibility that more sensitive measurements of somatic-motor control are possible, revealing some form of volitional control, which would render the diagnostic statement of CLIS at least for this case inaccurate.

We conclude that this BCI communication demonstration broke a “wall” of communication silence encircling this CLIS individual. This experience also highlights that individuals with extremely severe and incapacitating ailments for extended periods are capable of meaningful communication. The current BCI system has several limitations, as several software and hardware modifications would need to be implemented before the system could be used independently by the family or caretakers without the presence of a technically skilled team. The BCI-software is presently being modified to improve communication outcome and self-reliance of the family.

To conclude, this case study has demonstrated that a patient without any stable and reliable means of eye-movement control or identifiable communication route employed a neurofeedback strategy to modulate the firing rates of neurons in a paradigm allowing him to select letters to form words and sentences to express his desires and experiences.

### Dataset reported in this article

The dataset here spans days 106 to 247 after implantation. For the analysis of neurofeedback trials in Figure 3 and the corresponding main text, only blocks after day 123 were used because of a change in paradigm (before day 123, incorrect trials and time-outs were not differentiated). For Supplementary Figure 1, all neurofeedback blocks were used, as time-out trials were counted as ‘incorrect’ as well.

All speller sessions performed between days 106 and 247 were included in the analysis. Other experimental paradigms performed, such as motor attempt exploration or yes/no question are mentioned in summary (Figure 2), but not analysed.

## Data Availability

Firing rate data recorded in this period as well as log files are available upon request and freely after peer-reviewed publication.

## Acknowledgements

This research was supported by the Wyss Center for Bio and Neuroengineering, Geneva, Deutsche Forschungsgemeinschaft (DFG BI 195/77-1); German Ministry of Education and Research (BMBF) 16SV7701; CoMiCon, LUMINOUS-H2020-FETOPEN-2014-2015-RIA (686764), Bogenhausen Staedtische Klinik, Munich. The authors thank Andrew Jackson and Nick Ramsey for their comments on an earlier version of the manuscript. We thank the patient and his family.

## Author’s contribution

Ujwal Chaudhary – Initiation; Conceptualization; Ethics Approval; Performed 100% of the sessions with the patient before and after implantation; Neurofeedback paradigm; Manuscript writing.

Ioannis Vlachos – Software development, integration and testing; Data analysis; Neurofeedback paradigm implementation; Performed 10% of the sessions after the implantation.

Jonas B. Zimmermann – Neurofeedback paradigm implementation; Data analysis; Figures; Manuscript writing.

Arnau Espinosa – Software testing; EEG/EOG analysis; Figures.

Alessandro Tonin – Speller software development; Performed 30% of sessions before implantation and 15% of the sessions after implantation; EEG/EOG analysis; Figures.

Andres Jaramillo-Gonzalez – EEG/EOG analysis; Figures. Majid Khalili-Ardali – Graphical user interface development.

Helge R. Topka – Ethics Approval; Medical patient care; Clinical and diagnostic neurological procedures.

Jens Lehmberg – Ethics approval; Neurosurgery; Clinical care. Gerhard M. Friehs – Neurosurgical training.

Alain Woodtli – Ethics approval; BfArM approval; Neurosurgical training. John P. Donoghue – Initiation; Conceptualization, writing, review, editing.

Niels Birbaumer – Initiation; Conceptualization; Coordination, Clinical-psycholgical procedures and care; Neurofeedback paradigm; performed 30% of the sessions with UC; Ethics approval; BfArM approval; Manuscript writing.

## List of Supplementary Figures

**Supplementary Figure S1: Audio-Neurofeedback Accuracy** – Accuracy in all neurofeedback blocks from day 106 (first time neurofeedback speller was attempted) to day 247 (minimum/median/maximum number of blocks per day: 3/7/16). A) Each feedback block is represented as a dot. Daily maximum accuracy connected by black line. B) and C) feedback trials on two different blocks.

**Supplementary Figure S2: Electrooculogram (EOG) features** – evolution of the A) maximum, B) mean, C) variance, D) power in the band 0-2 Hz, and E) power in the band 2-4 Hz of the EOG signal for yes/no questions answered by the patient during the period March 2018 to November 2019. For each day, the vertical and horizontal EOG raw signal was filtered between 0.05Hz and 40Hz. Each yes and no trial was extracted and corrected by subtracting the mean of the trial’s baseline. Then the range of the amplitude of the EOG signal was calculated separately for each trial as the difference between the maximum and minimum of the signal. The figure shows the mean and the standard error of the mean of the extracted range of the amplitude of the vertical and horizontal EOG signal across each day for yes and no trials. For each day, a Mann-Whitney U-test was performed between yes and no results separately for the horizontal and vertical EOG represented by the significant p-values shown on the top of the bars. The table in the middle of the figure lists the date and the number of yes and no trials used for vertical and horizontal EOG. The x-axis represents the date of the sessions, and the y-axis represents the amplitude in microvolts. The vertical red line indicates the date of the implantation: 19 March 2019. The month of all the non-invasive sessions performed after implantation are highlighted in red on the x-axis

